# Polygenic risk scores and Parkinson’s disease in South Africa: Moving towards ancestry-informed disease prediction

**DOI:** 10.1101/2025.09.29.25336747

**Authors:** Kathryn Step, Carene Anne Alene Ndong Sima, Spencer Grant, Jonggeol Jeffrey Kim, Emily Waldo, Soraya Bardien, Ignacio F. Mata, the Global Parkinson’s Genetics Program (GP2)

**Author notes:** **Corresponding author** Ignacio F. Mata.

## Abstract

Parkinson’s disease (PD) is a complex neurodegenerative disorder with environmental and genetic influences. Using genotyping array data of 661 South African PD cases and 737 controls, a polygenic risk score (PRS) analysis was conducted using PRSice-2. Summary statistics were used from two PD association studies as base datasets. The target dataset was split into training (70%; n=979) and validation (30%; n=419) cohorts. Various clumping window sizes, linkage disequilibrium thresholds, and *p*-value thresholds were tested to determine the optimal combination for risk prediction. Additionally, we investigated the variance explained by different combinations of covariates. Finally, top contributing variants were identified and cross-referenced with inferred local ancestry to assess ancestry-specific effects. Overall, modest predictive performance was observed (AUC: 0.6307-0.6311). Age at recruitment was the strongest individual predictor, while sex contributed the least. Local ancestry analysis indicated that the top contributing variants were not ancestry-specific risk variants. These findings provide the first evaluation of PRS performance for PD in a highly admixed South African cohort, underscoring the importance of including underrepresented populations in genetic risk prediction.

## Introduction

Parkinson’s disease (PD) is a complex neurodegenerative disorder with a range of motor and non-motor symptoms (Lima et al. 2012). PD has the fastest-growing rates of prevalence, disability, and deaths among the neurological disorders (GBD 2021 Nervous System Disorders Collaborators 2024). By 2050, it is predicted that there will be 25.2 million people living with PD globally, with the highest increase in prevalence projected for West Africa (Su et al. 2025).

Originally, PD was viewed as a sporadic disease resulting from environmental exposures (Chen & Ritz 2018; Ascherio & Schwarzschild 2016). However, the past two and a half decades of PD research have revealed a complex disease etiology consisting of monogenic causes as well as gene-environment and gene-gene interactions (Chen & Ritz 2018). Based on previous studies, approximately 5-10% of PD cases are expected to be monogenic, the result of single-gene mutations with large effect sizes (Jia et al. 2022)h. These genes are identified through genetic linkage analysis in families with multiple affected members (Reed et al. 2019; Klein & Westenberger 2012). While monogenic forms of the disease are commonly observed in familial PD, they can occur in sporadic cases, often due to deleterious genetic variants with incomplete penetrance (Reed et al. 2019). Moreover, increasingly larger genome-wide association studies (GWAS) have demonstrated a polygenic contribution to sporadic PD, where multiple risk variants, each with small effect sizes, jointly increase PD risk (Lesage & Brice 2009). Through these studies, significant advances have been made in our understanding of defining disease risk through identifying variants with low risk (Nalls et al. 2019).

These risk variants, also known as susceptibility variants, can be used for risk prediction through polygenic risk scores (PRS) (Step et al. 2024). In a PRS analysis, the variants identified through GWAS, along with their effect sizes (whether conferring increased or decreased risk) are combined to estimate an individual’s genetic predisposition to a disease or trait (Ndong Sima et al. 2024). In 2016, the first report on polygenic risk and clinical outcomes for PD was published (Pihlstrøm et al. 2016). Since then, several studies have evaluated PRS for PD risk prediction, with predictive performance, assessed by the area under the receiver operating characteristic curve (AUC), ranging from approximately 60% to 76% (Li et al. 2019; Foo et al. 2020; Nalls et al. 2019; Ibanez et al. 2017; Loesch et al. 2022). This is largely dependent on the number of single-nucleotide polymorphisms (SNPs) included and the population characteristics (Dehestani et al. 2021).

The primary goal of PRS analysis is to classify individuals based on their relative risk for developing a disease. Ultimately, this could play a role in risk stratification, early intervention strategies, and tailored precision medicine approaches (Step et al. 2024; Dehestani et al. 2021). Here, we aimed to perform the first PRS analysis for PD in a South African study collection, assessing the predictive performance across two different base datasets to determine the optimal strategy for calculating PRS for a non-communicable disease such as PD in a multi-way admixed population, with a focus on predicting disease status.

## Methods

### Participant demographics

Study participants were recruited from 2002 until June 2020 (Health Research Ethics Committee, Stellenbosch University, 2002C/059), as part of the South African Parkinson’s Disease Study Collection (**Supplementary Table 1**) (van Rensburg et al. 2022). Individuals living with PD (n=691) were diagnosed in accordance with the Queen’s Square Brain Bank Criteria (Hughes et al. 1992). In total, 826 controls were recruited as part of the study collection through blood donor clinics of the Western Province Blood Transfusion Service (van Rensburg et al. 2022).

### Genotyping, imputation, quality control, and ancestry inference

Genotyping was completed through the Global Parkinson’s Genetics Program (Global Parkinson’s Genetics Program 2021) using the NeuroBooster Array (v1.0, Illumina, San Diego, CA) (Bandres-Ciga et al. 2024). QC was performed using PLINK v1.9 and v2.0 (Purcell et al. 2007; Chang et al. 2015), as previously described (Step et al. 2025). Imputation was performed using the TOPMed Imputation Server (Das et al. 2016). The related individuals (n=63) were identified using NAToRA (Leal et al. 2022) and a kinship coefficient of 0.0884 (second degree relation (Manichaikul et al. 2010)) and excluded from the analysis. After QC, 661 PD cases and 737 controls remained for downstream analysis (**Supplementary Table 1**). The South African population is five-way admixed (Swart et al. 2021), therefore, a reference panel was created using samples from the 1000 Genomes Project Phase III, including individuals of African (AFR), European (EUR), and South Asian (SAS) ancestries (Byrska-Bishop et al. 2022; Shriner et al. 2023). Additional individuals of Malaysian (MAL) ancestry and an indigenous hunter-gatherer Khoe-San (NAMA) population were included in the reference panel (Ragsdale et al. 2023), as previously described (Step et al. 2025). The reference files were phased using the TOPMed Imputation Server (Das et al. 2016). For the South African dataset, local ancestry inference was performed using G-Nomix (Hilmarsson et al. 2021), as previously described (Step et al. 2025) (**Supplementary Figure 1**).

### Polygenic risk score calculation

A standard PRS analysis includes the following main steps (**Figure 1**): (1) data quality control (QC) and preparation, (2) calculation of the PRS, and (3) assessment of PRS performance (Ndong Sima et al. 2024). A PRS analysis utilizes two independent datasets: a discovery dataset and a target dataset (Ndong Sima et al. 2024). The discovery dataset consists of GWAS summary statistics, including effect sizes for each variant. The target dataset contains individual-level genotype data, from which SNP dosages are derived for variants included in the PRS calculation. In general, PRS is computed for each individual as the sum of the dosages of risk alleles at selected SNPs, weighted by their corresponding effect sizes from the discovery dataset (Choi et al. 2020). For this study, we used PRSice-2 v2.3.3 (Choi & O’Reilly 2019), which implements a traditional clumping and thresholding approach to the PRS calculation.

**Figure 1:**
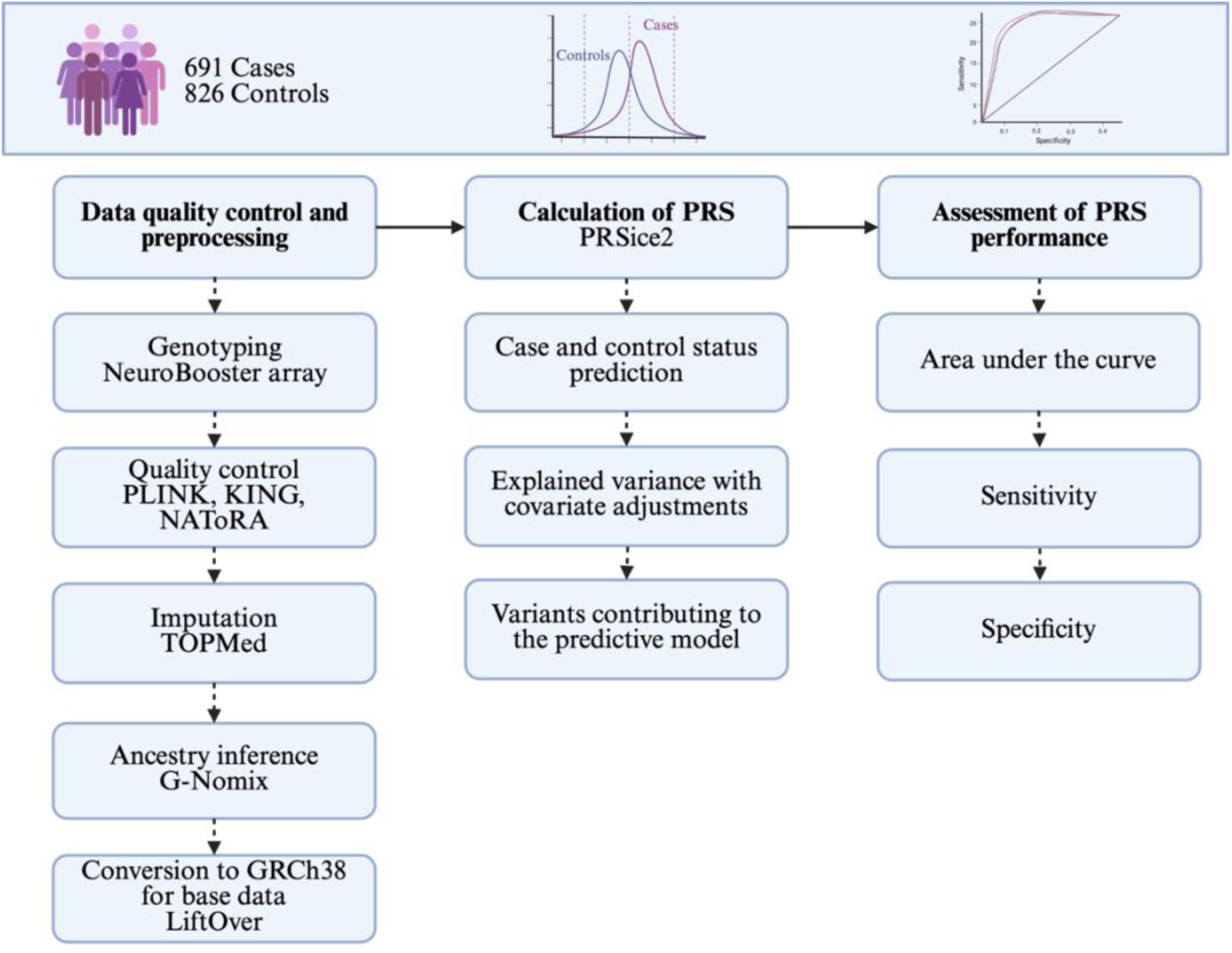
An overview of the methods followed in the present study. LiftOver was specific to the summary statistics used as the base datasets. PRS, polygenic risk score.

### Polygenic risk score: file preparation

The covariate file was created using the marginal segment probability output files from G-Nomix. These files contain ancestry inferences for the parental populations for each individual and both haplotypes across inferred local ancestry windows. The ancestry proportions were calculated by extracting the ancestry window information, specifically calculating the total genomic span of each parental ancestry and normalising the values to determine relative ancestry contribution per individual. In addition to the ancestry proportions, covariates included age and sex. Full summary statistics were obtained from the NHGRI-EBI GWAS catalog (Cerezo et al. 2025) on 05/09/2024 for studies GCST009325 (Nalls et al. 2019) and GCST90275127 (Kim et al. 2024). These two base datasets were used to assess and compare the predictive power of the EUR dataset relative to a multi-ancestry dataset which is better matched to the admixture of the South African cohort. This approach was used to evaluate whether ancestry-matched summary statistics enhanced predictive performance in an admixed population, as traditional EUR-derived GWAS does not fully capture the genetic architecture in diverse populations, like the South African population. The summary statistics, serving as the base dataset, were converted from GRCh37 to GRCh38 using LiftOver v1.0 (Perez et al. 2025). The South African Parkinson’s disease data, serving as the target dataset, was randomly split into two cohorts: 70% of the samples were in the training dataset (n=979 individuals; n=445 cases; n=534 controls), and 30% in the validation dataset (n=419 individuals; n=216 cases; n=203 controls). To assess the robustness of the data split, we ran the PRS analysis across 20 random splits and compared the distribution of the AUC values with the original split (**Supplementary Table 2**).

### Polygenic risk score analysis case status

The PRS analysis was run using PRSice-2 (Choi & O’Reilly 2019), which applies clumping and thresholding based on linkage disequilibrium (LD) and associated *p*-values. The optimal predictive PRS model is evaluated using Nagelkerke’s pseudo-R^2^ (R^2^) (Ndong Sima et al. 2024). Here, we tested a range of parameter combinations to determine the optimal model. For this analysis, clumping was conducted using window sizes of 100kb, 250kb, and 500kb; LD thresholds (r^2^) of 0.1, 0.2, 0.5, and 0.8; as well as the *p*-value thresholds of 1×10^-3^, 1×10^-5^, 1×10^-6^, and 5×10^-8^. All analyses were performed using logistic regression, adjusting for sex, age, and population structure covariates (AFR, EUR, NAMA, SAS). To prevent perfect multicollinearity, we excluded the MAL ancestry from the covariates, as it represented the smallest ancestral contribution. To obtain robust significance estimates, empirical *p*-values were calculated using 10,000 phenotype permutations. The initial search for optimal PRS parameters was conducted on 70% of the dataset (training cohort), and the best-performing parameters were then applied to fit a new model on the remaining 30% (validation cohort). The AUC was used to evaluate the performance of each PRS model using the pROC package (Robin et al. 2011) in R v4.2.0, providing a quantitative metric for comparing models (Konuma & Okada 2021). Additionally, predicted probabilities from a logistic regression model were converted to binary disease status using a fixed threshold of 0.5, and model performance was evaluated using accuracy, balanced accuracy, sensitivity, and specificity calculated at this threshold. Additionally, the positive predictive value and negative predictive values were calculated at multiple top-percentile thresholds (5%, 10%, and 20%) using the global PD prevalence of 1.386×10^-4^ (Ou et al. 2021).

### Assessment of explained variance based on covariate inclusions

In addition to identifying the optimal input parameters for disease prediction across both base datasets, we evaluated the contribution of different covariate combinations to the variance explained. Using PRSice-2 output, we examined the variance models across seven covariate inclusion scenarios, each incorporating a distinct combination of the following variables: age, sex, and local ancestry proportions. This analysis allowed us to quantify the incremental variance explained by each covariate and their combinations, providing insight into their individual and joint effects on disease risk prediction.

### Identification of the top variants contributing to the polygenic risk score

To identify the variants contributing most to the predictive performance of the PRS model, we used the PRSice-2 output file listing the SNPs included in the final model. New base datasets were generated by subsetting the original summary statistics to include only the model SNPs. The PRS analysis was systematically rerun, each time excluding a single SNP from this filtered base file while keeping the target constant. For each run, we evaluated the change in AUC to quantify the individual contribution of each SNP to the model’s predictive power, where the variants were ranked based on the resulting decrease in AUC relative to the original model that included all SNPs. The top contributing variants were then cross-referenced with inferred local ancestry windows to assess whether their predictive effects were ancestry-specific.

## Results

### Preprocessing for PRSice-2

The analysis included 35,075,375 variants in the two target datasets: a training dataset comprising 70% of the cohort (n=979) and a validation dataset comprising 30% (n=419). To identify the optimal PRS model, we conducted the analysis using two sets of summary statistics as the base data: Nalls *et al*., (2019) and Kim *et al*., (2024). For each, we evaluated varying combinations of clumping parameters, LD thresholds, clumping window sizes, and *p*-value thresholds for SNP inclusion (**Supplementary Figure 2 and 3**). The analyses were first performed on the training dataset to determine the optimal thresholds and then replicated in the validation dataset to evaluate predictive performance.

### Optimization and training of PRSice-2 for PD status prediction

One of the main uses of PRS is to predict case status according to their genetic risk or predisposition, making it a useful prognostic tool (**Supplementary Figure 4**) (Dehestani et al. 2021). Using the EUR-based summary statistics (Nalls et al. 2019), we tested 48 combinations of parameters to identify the best-performing PRS model for PD status prediction (**Supplementary Figure 5**). The highest predictive performance was observed under the following parameter set: 100kb clumping window, *r²* = 0.5, and a SNP inclusion threshold of *p*-value = 1×10^−3^. Here, the full R^2^ explained 37.11% of the variance in the disease phenotype (PRS R^2^ = 0.0168; adjusted R^2^ = 0.0176), with a strong association (*p*-value = 4.16×10^−5^; empirical *p*-value = 2.0×10^−4^). A total of 614 SNPs remained after clumping. The PRS analysis was conducted on the validation dataset (30%) using the parameter combination that demonstrated the highest predictive performance in the training dataset (100kb clumping window, *r²* = 0.5, *p*-value = 1×10^−3^). When applied to the validation set, the optimal model (defined at a stricter SNP inclusion threshold of *p*-value = 5×10^−5^) explained 35.60% of the variance in case-control status (PRS R² = 0.0374; adjusted R² = 0.0376; *p*-value = 9.1×10^−5^). The *p*-value remained robust after permutation testing (empirical *p*-value = 3.0×10^−4^). A total of 486 SNPs were included in this model.

Given the high genetic admixture in the South African cohort (Step et al. 2025), we evaluated predictive accuracy using multi-ancestry summary statistics from Kim *et al*. (2024). We tested the same 48 parameter combinations as for the previous analysis to identify the optimal PRS model (**Supplementary Figure 6**). The highest predictive performance was observed under the following parameter set: a 100kb clumping window, *r²* = 0.5, and a *p*-value threshold of 1×10^−3^. The full R^2^ explained 37.99% of the variance (PRS R² = 0.0251; adjusted R² = 0.0264; *p*-value = 4.74×10^−7^; empirical *p*-value = 1.0×10^−4^). A total of 651 variants remained after clumping in this model. Unlike the EUR-based analysis, the multi-ancestry summary statistics have shorter and more fragmented LD blocks, therefore more SNPs are retained after clumping in comparison to the EUR-based summary statistics. Using the multi-ancestry meta-analysis summary statistics from Kim *et al*. (2024) and the previously identified best threshold combination, we performed PRS analysis on the validation dataset (30%). The best-fitting model, defined by a *p*-value threshold of 1×10^−3^ and including 969 SNPs, explained 36.34% of the variance in disease phenotype (PRS R² = 0.0446), with an adjusted R² of 0.0450 after accounting for covariates. The PRS coefficient was 0.145 (SE = 0.034; *p*-value = 2.04×10^−5^; empirical *p*-value = 1.0×10^−4^).

### Assessment of model performance

The AUC, sensitivity, and specificity were assessed using the two base datasets as well as the training and validation cohorts of the target dataset (**Table 1**; **Figure 2**). Additionally, we assessed the mean AUC (Nall *et al* 2019: 0.6286 ± 0.013; Kim *et al* 2024: 0.6278 ± 0.1223) across 20 random data splits to assess the robustness of our data split into training and validation cohorts. The results were highly consistent with the original split presented below, indicating the predictive performance was stable and not notably influenced by the random split.

**Table 1:** Model performance across base and target datasets.

| **Base and target data** | **Nalls with training** | **Nalls with validation** | **Kim with training** | **Kim with validation** |
| --- | --- | --- | --- | --- |
| **Best p-value threshold** | 8.50E-05 | 5.00E-05 | 5.50E-05 | 1.55E-04 |
| **PRS.R2** | 0.0168 | 0.0374 | 0.0251 | 0.0446 |
| **PRS.R2.adj** | 0.0176 | 0.0376 | 0.0264 | 0.0450 |
| **Full.R2** | 0.3711 | 0.3560 | 0.3799 | 0.3634 |
| **Null.R2** | 0.3535 | 0.3184 | 0.3535 | 0.3184 |
| **Coefficient** | 0.0726 | 0.1138 | 0.1180 | 0.1450 |
| **Standard Error** | 0.0177 | 0.0291 | 0.0234 | 0.0340 |
| **P-value of model fit** | 4.16E-05 | 9.10E-05 | 4.74E-07 | 2.04E-05 |
| **Number of SNPs included** | 614 | 486 | 651 | 969 |
| **Empirical-P** | 2.00E-04 | 3.00E-04 | 1.00E-04 | 1.00E-04 |
| **AUC (95% CI)** | 0.6307<br>(0.5955-0.6659) | 0.6290<br>(0.5760-0.6821) | 0.6311<br>(0.5961-0.6661) | 0.6206<br>(0.5672-0.6741) |
| **Accuracy (95% CI)** | 0.6139<br>(0.5826-0.6445) | 0.5895<br>(0.5407-0.6370) | 0.6108<br>(0.5795-0.6415) | 0.5609<br>(0.5119-0.6090) |
| **Balanced Accuracy** | 0.5993 | 0.5873 | 0.5970 | 0.5590 |
| **Sensitivity** | 0.4382 | 0.6574 | 0.4400 | 0.6204 |
| **Specificity** | 0.7603 | 0.5172 | 0.7528 | 0.4975 |
| Legend: The DeLong p-value for the training dataset is 0.9778 and for the validation dataset it is 0.7057, showing the base dataset does not influence the model. Accuracy, balanced accuracy, sensitivity, and specificity were calculated using a 0.5 threshold. AUC, area under the curve; CI, confidence interval; PRS R2 is the variance explained by the PRS. PRS R2 adjusted is the PRS R2 adjusted for ascertainment. The Full R2 is the variance explained by the full model (including the covariates). Null R2 is the variance explained by the covariates only |  |  |  |  |

**Figure 2:**
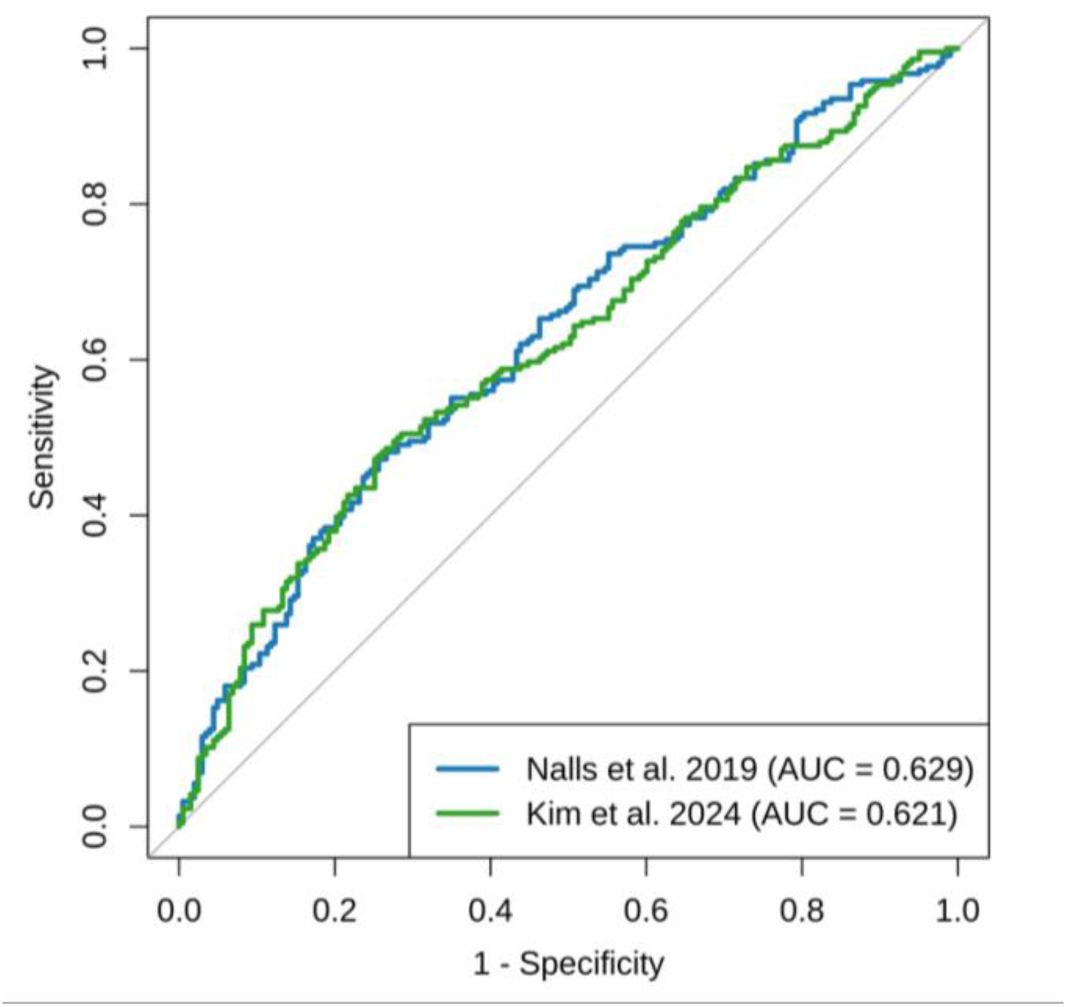
Receiver Operating Characteristic (ROC) Curve Comparing Polygenic Risk Score (PRS) Models for Case Status. The ROC curves compare the predictive performance of two PRS models for Parkinson’s disease: one based on Nalls et al. (2019) (blue) and the other on Kim et al. (2024) (green). The area under the curve (AUC) indicates the discriminative ability of each model, with higher AUC values reflecting better classification of cases and controls. The AUC for the Nalls model is 0.629, while the AUC for the Kim model is 0.621. PRS were calculated and matched to phenotype data from the same sample set (N = 419).

Using the Nalls *et al*., 2019 summary statistics, the PRS demonstrated a moderate ability to distinguish between PD cases and controls, with an AUC of 0.6307 (95% CI: 0.596-0.666) in the training dataset and 0.6290 (95% CI: 0.576-0.682) in the validation dataset. At a fixed probability threshold of 0.5, classification accuracy was 61.39% (95% CI: 0.583-0.645) in the training dataset and 58.95% (95% CI: 0.541-0.637) in the validation dataset. The observed balanced accuracy was similar between datasets (59.93% and 58.73%, respectively). Sensitivity was higher in the validation dataset (65.74%) compared to the training dataset (43.82%), whereas specificity was higher in the training dataset (76.03% versus 51.72%).

In contrast, the PRS model constructed using summary statistics from Kim *et al*., 2024 yielded a comparable discriminative ability, with an AUC of 0.6311 (95% CI: 0.596-0.666) in the training dataset and 0.6206 (95% CI: 0.5672-0.6741) in the validation dataset. Classification accuracy at the 0.5 threshold was 61.08% (95% CI: 0.580-0.642) and 56.09% (95% CI: 0.512-0.609) in the training and validation datasets, respectively. Balanced accuracy was modest in both datasets (59.70% and 55.90%), with sensitivity again higher in the validation dataset (62.04%) compared to the training dataset (44.00%). Specificity followed the same pattern as observed in the EUR-based base dataset (Nalls et al. 2019), with higher values in the training dataset (75.28%) relative to validation (49.75%).

Finally, we evaluated the predictive performance of the PRS using top percentile thresholds (**Supplementary Table 3**). The sensitivity increased as more cases were included when the threshold was lowered, while specificity decreased correspondingly. The positive predictive value remained low across all thresholds, whereas the negative predictive values were consistently high (>99%). The overall patterns observed were similar between the PRS derived from the two base datasets with minor differences in the number of cases captured at the top 5% threshold.

These results suggest that while the PRS performs moderately overall, its predictive ability varies across datasets. This may be due, in part, to the slight imbalance of cases and controls, particularly in the training cohort, which biases the model toward predicting controls and results in a higher sensitivity but lower specificity.

### Covariate contribution to the variance observed

We evaluated the contribution of covariates to the explained variance by examining their effect on model performance (**Table 2**, **Supplementary Table 4**). We looked at the three variance models (PRS R^2^, Full R^2^, and Null R^2^) under seven different covariate inclusion scenarios, using each possible combination of covariates: age, sex, and local ancestry proportions.

**Table 2:** Variance explained by polygenic risk score models with different covariate adjustments for the training dataset.

| Study | PRS R<sup>2</sup> | PRS R<sup>2</sup> adjusted | Full R<sup>2</sup> | Null R<sup>2</sup> | Number of SNPs | Empirical p-value | Covariates included |
| --- | --- | --- | --- | --- | --- | --- | --- |
| Nalls | 0.017 | 0.018 | 0.371 | 0.354 | 614 | 2.00E-04 | AGE, SEX, ANC |
| Nalls | 0.039 | 0.039 | 0.320 | 0.281 | 1838 | 1.00E-04 | AGE |
| Nalls | 0.057 | 0.047 | 0.054 | 0.007 | 658 | 1.00E-04 | SEX |
| Nalls | 0.039 | 0.033 | 0.089 | 0.056 | 564 | 1.00E-04 | ANC |
| Nalls | 0.038 | 0.038 | 0.330 | 0.292 | 1838 | 1.00E-04 | AGE, SEX |
| Nalls | 0.016 | 0.017 | 0.358 | 0.341 | 614 | 4.00E-04 | AGE, ANC |
| Nalls | 0.040 | 0.034 | 0.098 | 0.064 | 564 | 1.00E-04 | SEX, ANC |
| Kim | 0.025 | 0.026 | 0.380 | 0.354 | 651 | 1.00E-04 | AGE, SEX, ANC |
| Kim | 0.035 | 0.035 | 0.316 | 0.281 | 1480 | 1.00E-04 | AGE |
| Kim | 0.067 | 0.054 | 0.061 | 0.007 | 1376 | 1.00E-04 | SEX |
| Kim | 0.055 | 0.046 | 0.103 | 0.056 | 867 | 1.00E-04 | ANC |
| Kim | 0.037 | 0.037 | 0.329 | 0.292 | 1480 | 1.00E-04 | AGE, SEX |
| Kim | 0.023 | 0.024 | 0.366 | 0.341 | 651 | 1.00E-04 | AGE, ANC |
| Kim | 0.058 | 0.049 | 0.113 | 0.064 | 1330 | 1.00E-04 | Sex, ANC |

Legend: Table shows PRS R<sup>2</sup> values, adjusted and unadjusted, with corresponding full and null model R<sup>2</sup>, empirical significance, and number of SNPs included, stratified by covariates used in each model. PRS R<sup>2</sup> is the variance explained by the PRS. PRS R<sup>2</sup> adjusted is the PRS R<sup>2</sup> adjusted for ascertainment. The Full R<sup>2</sup> is the variance explained by the full model (including the covariates). Null R<sup>2</sup> is the variance explained by the covariates only. ANC, Ancestries; SNPs, Single nucleotide polymorphisms

We assessed this using the Nalls *et al*., 2019 summary statistics as the base data. Here, the highest PRS R², representing the variance explained by the PRS after accounting for covariates, was observed when adjusting for sex only (PRS R^2^, = 0.057; adjusted R^2^ = 0.047). However, this model had the lowest full model R² (0.054) and the largest drop from the null model (0.007), suggesting limited overall model performance despite a high PRS-specific contribution. Additionally, the highest Null R^2^, reflecting variance explained by covariates alone, was observed when including age, sex, and ancestries (Null R^2^ = 0.354), indicating this combination contributed most to explaining outcome variability independent of genetic risk. In contrast, the lowest Null R² was seen in the model including only sex, suggesting this covariate alone has a limited contribution to the model variance and cannot be used alone to predict risk. Moreover, the model adjusting for age, sex, and ancestries explained a substantial proportion of the variance in the full model (Full R^2^ = 0.371), with a modest PRS R^2^ of 0.017 and a significant empirical *p*-value (*p*-value = 2.0×10^−4^). Models that included age and ancestry, or sex and ancestry, had similarly low PRS R^2^ values (∼0.017 to 0.034) but retained significant *p*-values (*p*-values ≤ 1.0×10^−4^), reflecting a consistent but modest PRS contribution across covariate combinations. All models remained statistically significant based on empirical *p*-values (range: 1.0×10^−4^ to 4.0×10^−4^), although the magnitude of variance explained by genetic and covariate components varied depending on the covariate combination.

For Kim *et al*., 2024, the same seven covariate combinations were assessed. The highest PRS R^2^, showing the variance explained by the PRS model only, was observed when adjusting for sex (PRS R^2^ = 0.067), while the lowest was seen when adjusting for age and ancestries (PRS R^2^ = 0.023). The Null R^2^ values are consistent with those observed in the EUR-based summary statistics, as expected, since they capture the variance explained by covariates alone and are independent of genetic influence. Finally, all models were statistically significant (empirical *p*-values: ∼1.0×10^−4^), and the variance explained by the PRS was dependent on the covariate structure, highlighting the impact of covariate selection on model performance.

We evaluated whether the inclusion of PRS improved the predictive performance for the models including only covariates (**Table 3**). For this, we looked at both base dataset summary statistics as well as the training and validation cohorts. The addition of the PRS consistently increased the AUC for all covariate combinations. The improvement was more pronounced in models with fewer covariates showing statistical significance (DeLong *p*-value < 0.05). For models including the full set of covariates (age, sex, and ancestral components), the PRS still increased the AUC, though the change was smaller and in some cases not statistically significant (DeLong *p*-value > 0.05). These results indicate that the PRS provides a meaningful addition to the predictive power beyond the covariates included.

**Table 3:** Effect of adding the polygenic risk score on predictive performance of covariate models.

| Covariates | AUC (Covariates only) | AUC (Full model: Covariates + PRS) | DeLong p-value |
| --- | --- | --- | --- |
| **Training: Nalls et al 2019** |  |  |  |
| AGE | 0.7645 | 0.7922 | 4.33E-05 |
| SEX | 0.5403 | 0.6365 | 4.29E-07 |
| AGE+SEX | 0.7700 | 0.7949 | 1.10E-04 |
| AGE+AFR+ANC | 0.8145 | 0.8196 | 0.1846 |
| SEX+ANC | 0.6364 | 0.6703 | 0.0065 |
| ANC | 0.6218 | 0.6604 | 0.0049 |
| AGE+SEX+ANC | 0.8159 | 0.8226 | 0.0824 |
| **Validation: Nalls et al 2019** |  |  |  |
| AGE | 0.7511 | 0.7923 | 0.0033 |
| SEX | 0.5863 | 0.6639 | 3.27E-04 |
| AGE+SEX | 0.7688 | 0.7994 | 0.0126 |
| AGE+AFR+ANC | 0.7909 | 0.8090 | 0.0770 |
| SEX+ANC | 0.6399 | 0.6865 | 0.0069 |
| ANC | 0.5942 | 0.6574 | 0.0093 |
| AGE+SEX+ANC | 0.7996 | 0.8160 | 0.0929 |
| **Testing: Kim et al 2024** |  |  |  |
| AGE | 0.7645 | 0.7863 | 2.74E-04 |
| SEX | 0.5403 | 0.6392 | 3.45E-07 |
| AGE+SEX | 0.7700 | 0.7910 | 4.21E-04 |
| AGE+AFR+ANC | 0.8145 | 0.8227 | 0.0649 |
| SEX+ANC | 0.6364 | 0.6831 | 7.25E-04 |
| ANC | 0.6218 | 0.6723 | 6.25E-04 |
| AGE+SEX+ANC | 0.8159 | 0.8258 | 0.0329 |
| **Validation: Kim et al 2024** |  |  |  |
| AGE | 0.7511 | 0.7868 | 0.0051 |
| SEX | 0.5863 | 0.6498 | 0.0030 |
| AGE+SEX | 0.7688 | 0.7987 | 0.0103 |
| AGE+AFR+ANC | 0.7909 | 0.8124 | 0.0552 |
| SEX+ANC | 0.6399 | 0.6825 | 0.0203 |
| ANC | 0.5942 | 0.6650 | 0.0051 |
| AGE+SEX+ANC | 0.7996 | 0.8199 | 0.0538 |
| Legend: Comparison of model discrimination with and without the polygenic risk score (PRS). For each set of covariates, we report the area under the ROC curve (AUC) for the covariates-only model and the full model including the PRS, along with the p-value from the DeLong test. Significant p-values indicate that inclusion of the PRS significantly improves model discrimination beyond the covariates alone. AFR, African; ANC, Ancestral components; EUR, European; NAMA, Nama; SAS, South Asian |  |  |  |

### Analysis of top contributing variants reveals generalised population disease risk

To identify variants contributing most to the predictive accuracy of the model, we assessed their individual impact across the two base datasets and the training cohort, and aligned these findings with the inferred local ancestry of variant carriers (**Table 4**). In the Nalls *et al*., 2019 base dataset, rs78231164 nearest to *RANBP2* and *EDAR* emerged as the top contributing SNP to the predictive model, with its exclusion resulting in the greatest reduction in AUC (ΔAUC = -0.0025). Among the variant carriers, 76.84% had inferred AFR ancestry, consistent with gnomAD v.4.1.0, where this variant is more prevalent in AFR populations (Chen et al. 2024). The cumulative difference in AUC between the first- and tenth-ranked top contributing variants was -0.0013, indicating a low impact of individual variants on model performance. For the Kim *et al*., 2024 summary statistics as the base dataset, the top variant contributing to the model was rs4889571 nearest to *FBXL19* (ΔAUC = -0.0022). Among variant carriers in our cohort, 76.82% were of EUR ancestry and 9.95% were AFR, whereas in gnomAD v4.1.0, this variant is most frequent in EAS populations (Chen et al. 2024), reflecting differences in population-specific allele frequencies. For the top ten variants, the cumulative difference in AUC was -0.001, similar to what was observed in the Nall *et al*. 2019 base dataset. In terms of the inferred local ancestry, the top contributing SNPs were distributed across the five ancestral components observed in the South African study cohort. However, the largest percentages of variant carriers were seen in the AFR and EUR ancestral components, which is expected given that these ancestries make up the largest proportions of the study population (Step et al. 2025).

**Table 4:**
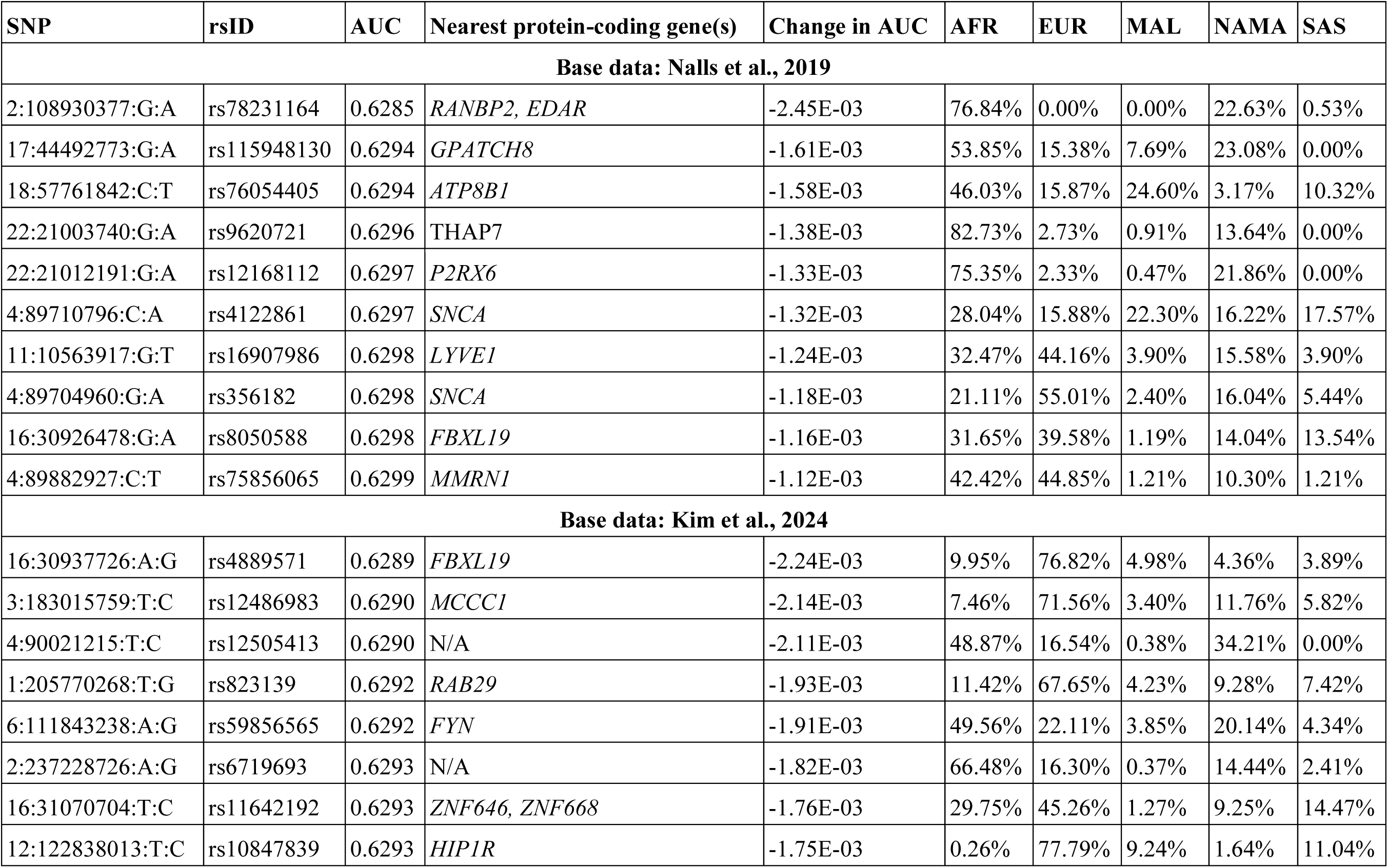
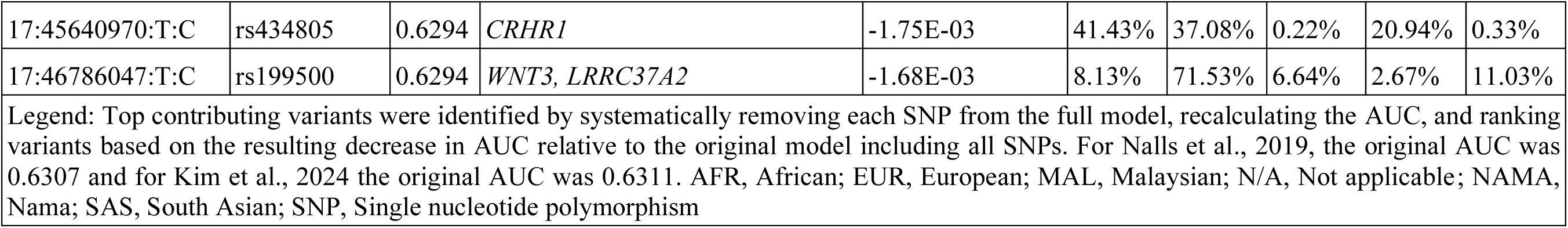
The top variants contributing to the polygenic risk score model for Parkinson’s disease case control status in the South African cohort for two base datasets.

## Discussion

To our knowledge, this is the first study to evaluate PRS for PD prediction in a South African cohort. We used a well-established PRS software, PRSice-2, and leveraged summary statistics from both EUR-ancestry and multi-ancestry GWAS for PD. Despite the summary statistics not fully matching the genetic background of our cohort, which is five-way admixed (Step et al. 2025), our findings demonstrate that polygenic models can still capture a modest but significant proportion of the variance in PD susceptibility. This highlights the utility of PRS in diverse and underrepresented populations.

Using PRSice-2, we identified the best parameter sets for both the Nalls *et al*. (2019) and Kim *et al*. (2024) summary statistics. In the training dataset, we found that clumping parameters (e.g., r² and window size) and *p*-value thresholds had notable effects on predictive power. The optimal PRS derived from the Nalls *et al*. (2019) summary statistics achieved a full R^2^ of 37.11% in disease phenotype in the training set and 35.60% in the validation dataset, with an AUC of 0.6307. Similarly, using the Kim *et al*. (2024) summary statistics with more diverse populations, the best-performing PRS model had a full R^2^ of 37.99% in the training dataset and 36.34% in the validation dataset, with an AUC of 0.6311. Although the PRS derived from Kim *et al*. (2024) explained more variance in disease risk, its slightly higher AUC compared to Nalls *et al*. (2019) highlights how R² and AUC capture different aspects of predictive performance, the underlying genetic predisposition versus classification accuracy, respectively. This may suggest that multi-ancestry meta-analysis PRS better models genetic predisposition for distinguishing cases from controls in the study population. However, the observed AUC is similar to those previously reported for PD PRS which range from 0.620 to 0.760 (Dehestani et al. 2021).

Notably, the models that achieved the highest predictive accuracy tended to favor moderate clumping thresholds (r² = 0.5) and relatively lenient SNP inclusion thresholds (*p*-value ≤ 1×10^−3^), likely balancing the trade-off between including informative variants and controlling for LD-induced noise. Moreover, the validation of model performance in an independent subset of the cohort strengthens the robustness of these findings and supports the potential clinical utility of PRS in diverse populations.

Overall, our results highlight that while sex contributes modestly to the explained variance, accounting for approximately 3-7% across models, it cannot be used in isolation to predict disease status (Dehestani et al. 2021), as evidenced by the low full model R^2^ values when sex was the only covariate. In contrast, age appears to provide a more meaningful contribution, particularly when combined with other covariates such as ancestry. Importantly, the PRS R^2^ values were consistently low across all models and datasets, suggesting that genetic risk alone, as captured by current PRS methods, is insufficient for reliable disease prediction (Staerk et al. 2024). However, the DeLong test demonstrated that adding PRS to models with sex or ancestry alone can significantly improve AUC, whereas the incremental gain was not significant when age and ancestry covariates were already included. This underscores that the predictive contribution of PRS is dependent on demographic and ancestral factors. These findings underscore the importance of incorporating non-genetic covariates to enhance predictive performance, as previously illustrated in a PD context (Nalls et al. 2015). Ultimately, the limited variance explained by PRS alone constrains its current clinical utility for PD and emphasizes the need for integrative models that include both genetic and phenotypic information (Step et al. 2024).

The additional sensitivity and specificity analyses support the overall performance of our PRS models, showing results that are consistent with previous work on multi-ancestry populations (Saffie Awad et al. 2024). Our AUC values ranged from 0.6307 to 0.6311 across base datasets, with corresponding balanced accuracy values between 0.5993 and 0.5970. These values are comparable to those reported by Saffie Awad *et al*. (2024), whose AUC ranged from 0.505 to 0.651 across four ancestries. Our model performs similarly to theirs, particularly in populations with higher predictive power, such as the EUR populations. Our sensitivity and specificity values also reflect expected trade-offs: for example, when sensitivity increased (e.g., 0.6574), specificity tended to decrease (e.g., 0.5172), consistent with typical classification dynamics. Together, these findings reinforce the idea that PRS models retain moderate predictive power in diverse populations, but that further optimization may be needed to improve performance, especially in underrepresented groups. This aligns with previous reports showing PRS accuracies as low as 20-40% in AFR ancestries when EUR-based GWAS summary statistics are used as base datasets (Wang et al. 2022).

Moreover, we investigated whether the top contributing variants to the models are ancestry specific. The observed change in AUC between the top variants was minimal, which is expected given the small proportion of variance explained by genetic factors in the model. For both base datasets, the local ancestry window containing the variant of interest was predominantly of EUR and AFR ancestry. However, carriers of the variant were distributed across multiple ancestral backgrounds. This suggests that the top associated variants represent general disease risk factors rather than ancestry-specific signals.

Despite the moderate predictive performance observed, our study underscores both the promise and limitations of current PRS models in underrepresented populations. A key limitation of this study is the small sample size and the lower mean age of our control group relative to the case group. Moveover, the AUC values achieved reflect modest discriminative power. This finding suggests that while PRS can contribute to risk stratification, they are not yet sufficient for clinical decision-making on their own. Future studies incorporating ancestry-specific GWAS, functional annotations, and integrative risk models may further improve PRS accuracy in AFR and admixed populations. A further limitation for the study is the limited sample size and subsequent lack of an appropriate validation cohort.

A key strength of the study is its novelty, representing the first evaluation of PRS for PD in a South African study collection, thereby addressing a critical gap in genetic risk research. By including both EUR-based and multi-ancestry summary statistics, we were able to compare the PRS transferability across ancestries and assess how base dataset ancestral composition influences predictive performance. Furthermore, the systematic evaluation of clumping thresholds, LD parameter, and *p*-value thresholds to identify the optimal input parameter combinations further strengthens the methodological approach of this analysis. The incorporation of local ancestry inference allowed us to explore ancestry-specific or general risk factors with a finer resolution, providing biological context that extends beyond prediction alone. Finally, the application of PRS across various diseases, including diabetes and cancer (Mavaddat et al. 2019; Tremblay et al. 2021), has proven valuable for stratifying individuals at highest risk, rather than serving as a direct predictor of disease development (Ayoub et al. 2022). In this context, our study contributes by refining PD risk prediction for a smaller subset of individuals most at risk for developing PD.

In conclusion, our results highlight the importance of including diverse ancestral cohorts and relevant covariates when constructing and evaluating PRS models. By systematically assessing the variance observed across different covariate combinations, we highlight the contributions of demographic and genetic factors to disease risk prediction. Future efforts should continue to refine ancestry specific risk-models to ensure equitable translation of PRS from research into clinical applications for early screening, disease risk prediction, and precision medicine (Roberts et al. 2019).

## Data and code availability

Data used in the preparation of this article were obtained from the Global Parkinson’s Genetics Program (GP2; https://gp2.org). Specifically, we used Tier 2 data from GP2 release 9 (https://doi.org/10.5281/zenodo.14510099). GP2 data are available on AMP PD (https://amp-pd.org). For the NAMA dataset, the data analyzed in this study is subjected to the following licenses/restrictions: No new genetic data was generated for this study; however, summary statistics for the quality and accuracy assessment of the genetic data for the NAMA participants will be made available to researchers who meet the criteria for access after application to the Health Research Ethics Committee of Stellenbosch University. Requests to access the Nama datasets should be directed to Prof. Marlo Moller. Summary statistics for the base datasets are available from the NHGRI-EBI GWAS catalog (https://www.ebi.ac.uk/gwas/) accession numbers GCST009325 and GCST90275127. The QC and ancestry inference pipelines were developed and are maintained by Dr. Thiago Peixoto Leal and are available at https://github.com/MataLabCCF. Additionally, an overview of the analysis and any additional scripts not available through the Mata Lab GitHub can be found in the GP2 public domain on GitHub (https://github.com/GP2code/SouthAfrican_PD_PRS) and were given a persistent identifier via Zenodo (https://doi.org/10.5281/zenodo.16583859).

## Author contributions

Conceptualization: K.S., I.F.M., S.B. Data analysis and investigation: K.S., C.A.A.N.S., E.W. Writing: K.S. Reviewing and editing: K.S., C.A.A.N.S., E.W., S.G., J.J.K., I.F.M., S.B.

## Acknowledgements

We would like to acknowledge and thank the study participants for their contribution. This project was supported by the Global Parkinson’s Genetics Program (GP2; https://gp2.org). GP2 is funded by the Aligning Science Across Parkinson’s (ASAP) initiative and implemented by The Michael J. Fox Foundation for Parkinson’s Research (MJFF). For a complete list of GP2 members see https://doi.org/10.5281/zenodo.7904831. For the purpose of Open Access, the author has applied a CC BY public copyright licence to any Author Accepted Manuscript version arising from this submission. All figures were created using BioRender (https://www.biorender.com/). We would like to acknowledge Lim Shen-Yang, Tan Ai-Huey, and Azlina Ahmad-Annuar for their efforts in recruiting study participants from Malaysia. We thank Kate Andersh, Laurel Screven, and Kim Paquette for their work as scientific project managers for this project. We would like to acknowledge Dr. Thiago Peixoto Leal for his work in developing the scripts used in the analysis. We also acknowledge the Centre for High Performance Computing (CHPC), South Africa, for providing computational resources. *For open access, the author has applied a CC BY public copyright license to all Author Accepted Manuscripts arising from this submission*.

## References

Ascherio, A. & Schwarzschild, M.A., 2016. The epidemiology of Parkinson’s disease: risk factors and prevention. Lancet neurology, 15(12), pp.1257–1272.

Ayoub, A. et al., 2022. Polygenic risk scores: improving the prediction of future disease or added complexity? The British journal of general practice: the journal of the Royal College of General Practitioners, 72(721), pp.396–398.

Bandres-Ciga, S. et al., 2024. NeuroBooster array: A genome-wide genotyping platform to study neurological disorders across diverse populations. Movement disorders: official journal of the Movement Disorder Society, 39(11), pp.2039–2048.

Byrska-Bishop, M. et al., 2022. High-coverage whole-genome sequencing of the expanded 1000 Genomes Project cohort including 602 trios. Cell, 185(18), pp.3426–3440.e19.

Cerezo, M. et al., 2025. The NHGRI-EBI GWAS Catalog: standards for reusability, sustainability and diversity. Nucleic acids research, 53(D1), pp.D998–D1005.

Chang, C.C. et al., 2015. Second-generation PLINK: rising to the challenge of larger and richer datasets. GigaScience, 4, p.7.

Chen, H. & Ritz, B., 2018. The search for environmental causes of Parkinson’s disease: Moving forward. Journal of Parkinson’s disease, 8(s1), pp.S9–S17.

Chen, S. et al., 2024. A genomic mutational constraint map using variation in 76,156 human genomes. Nature, 625(7993), pp.92–100.

Choi, S.W., Mak, T.S.-H. & O’Reilly, P.F., 2020. Tutorial: a guide to performing polygenic risk score analyses. Nature protocols, 15(9), pp.2759–2772.

Choi, S.W. & O’Reilly, P.F., 2019. PRSice-2: Polygenic Risk Score software for biobank-scale data. GigaScience, 8(7). Available at: 10.1093/gigascience/giz082.

Das, S. et al., 2016. Next-generation genotype imputation service and methods. Nature genetics, 48(10), pp.1284–1287.

Dehestani, M., Liu, H. & Gasser, T., 2021. Polygenic risk scores contribute to personalized medicine of Parkinson’s disease. Journal of personalized medicine, 11(10), p.1030.

Foo, J.N. et al., 2020. Identification of Risk Loci for Parkinson Disease in Asians and Comparison of Risk Between Asians and Europeans: A Genome-Wide Association Study. JAMA neurology, 77(6), pp.746–754.

GBD 2021 Nervous System Disorders Collaborators, 2024. Global, regional, and national burden of disorders affecting the nervous system, 1990-2021: a systematic analysis for the Global Burden of Disease Study 2021. Lancet neurology, 23(4), pp.344–381.

Global Parkinson’s Genetics Program, 2021. GP2: The global Parkinson’s genetics program. Movement disorders: official journal of the Movement Disorder Society, 36(4), pp.842–851.

Hilmarsson, H. et al., 2021. High resolution ancestry deconvolution for next generation genomic data. bioRxiv. Available at: 10.1101/2021.09.19.460980.

Hughes, A.J. et al., 1992. Accuracy of clinical diagnosis of idiopathic Parkinson’s disease: a clinico-pathological study of 100 cases. Journal of neurology, neurosurgery, and psychiatry, 55(3), pp.181–184.

Ibanez, L. et al., 2017. Parkinson disease polygenic risk score is associated with Parkinson disease status and age at onset but not with alpha-synuclein cerebrospinal fluid levels. BMC neurology, 17(1), p.198.

Jia, F., Fellner, A. & Kumar, K.R., 2022. Monogenic Parkinson’s Disease: Genotype, Phenotype, Pathophysiology, and Genetic Testing. Genes, 13(3). Available at: 10.3390/genes13030471.

Kim, J.J. et al., 2024. Multi-ancestry genome-wide association meta-analysis of Parkinson’s disease. Nature genetics, 56(1), pp.27–36.

Klein, C. & Westenberger, A., 2012. Genetics of Parkinson’s disease. Cold Spring Harbor perspectives in medicine, 2(1), p.a008888.

Konuma, T. & Okada, Y., 2021. Statistical genetics and polygenic risk score for precision medicine. Inflammation and regeneration, 41(1), p.18.

Leal, T.P. et al., 2022. NAToRA, a relatedness-pruning method to minimize the loss of dataset size in genetic and omics analyses. Computational and structural biotechnology journal, 20, pp.1821–1828.

Lesage, S. & Brice, A., 2009. Parkinson’s disease: from monogenic forms to genetic susceptibility factors. Human molecular genetics, 18(R1), pp.R48–59.

Lima, M.M.S. et al., 2012. Motor and non-motor features of Parkinson’s disease - a review of clinical and experimental studies. CNS & neurological disorders drug targets, 11(4), pp.439–449.

Li, W.-W. et al., 2019. Association of the polygenic risk score with the incidence risk of Parkinson’s disease and cerebrospinal fluid α-synuclein in a Chinese cohort. Neurotoxicity research, 36(3), pp.515–522.

Loesch, D.P. et al., 2022. Polygenic risk prediction and SNCA haplotype analysis in a Latino Parkinson’s disease cohort. Parkinsonism & related disorders, 102, pp.7–15.

Manichaikul, A. et al., 2010. Robust relationship inference in genome-wide association studies. Bioinformatics (Oxford, England), 26(22), pp.2867–2873.

Mavaddat, N. et al., 2019. Polygenic risk scores for prediction of breast cancer and breast cancer subtypes. The American Journal of Human Genetics, 104(1), pp.21–34.

Nalls, M.A. et al., 2015. Diagnosis of Parkinson’s disease on the basis of clinical and genetic classification: a population-based modelling study. Lancet neurology, 14(10), pp.1002–1009.

Nalls, M.A. et al., 2019. Identification of novel risk loci, causal insights, and heritable risk for Parkinson’s disease: a meta-analysis of genome-wide association studies. Lancet neurology, 18(12), pp.1091– 1102.

Ndong Sima, C.A.A., et al., 2024. Methodologies underpinning polygenic risk scores estimation: a comprehensive overview. Human genetics, 143(11), pp.1265–1280.

Ou, Z. et al., 2021. Global trends in the incidence, prevalence, and years lived with disability of Parkinson’s disease in 204 countries/territories from 1990 to 2019. Frontiers in public health, 9, p.776847.

Perez, G. et al., 2025. The UCSC Genome Browser database: 2025 update. Nucleic acids research, 53(D1), pp.D1243–D1249.

Pihlstrøm, L. et al., 2016. A cumulative genetic risk score predicts progression in Parkinson’s disease: Genetic Risk and Progression in Parkinson’s Disease. Movement disorders: official journal of the Movement Disorder Society, 31(4), pp.487–490.

Purcell, S. et al., 2007. PLINK: a tool set for whole-genome association and population-based linkage analyses. American journal of human genetics, 81(3), pp.559–575.

Ragsdale, A.P. et al., 2023. Publisher Correction: A weakly structured stem for human origins in Africa. Nature, 620(7972), p.E11.

Reed, X. et al., 2019. The role of monogenic genes in idiopathic Parkinson’s disease. Neurobiology of disease, 124, pp.230–239.

van Rensburg, Z.J. et al., 2022. The South African Parkinson’s Disease Study Collection. Movement disorders: official journal of the Movement Disorder Society, 37(1), pp.230–232.

Roberts, M.C., Khoury, M.J. & Mensah, G.A., 2019. Perspective: The clinical use of polygenic risk scores: Race, ethnicity, and health disparities. Ethnicity & disease, 29(3), pp.513–516.

Robin, X. et al., 2011. pROC: an open-source package for R and S+ to analyze and compare ROC curves. BMC bioinformatics, 12(1), p.77.

Saffie Awad, P. et al., 2024. Insights into ancestral diversity in Parkinsons disease risk: A comparative assessment of polygenic risk scores. medRxiv: the preprint server for health sciences. Available at: https://www.medrxiv.org/content/10.1101/2023.11.28.23299090v1.

Shriner, D. et al., 2023. Universal genome-wide association studies: Powerful joint ancestry and association testing. HGG advances, 4(4), p.100235.

Staerk, C. et al., 2024. Generalizability of polygenic prediction models: how is the R2 defined on test data? BMC medical genomics, 17(1), p.132.

Step, K. et al., 2024. Exploring the role of underrepresented populations in polygenic risk scores for neurodegenerative disease risk prediction. Frontiers in neuroscience, 18. Available at: https://www.frontiersin.org/articles/10.3389/fnins.2024.1380860.

Step, K. et al., 2025. Genome-wide association analyses reveal susceptibility variants linked to Parkinson’s disease in the South African population using inferred global and local ancestry. medRxiv. Available at: 10.1101/2025.08.01.25331910.

Su, D. et al., 2025. Projections for prevalence of Parkinson’s disease and its driving factors in 195 countries and territories to 2050: modelling study of Global Burden of Disease Study 2021. BMJ (Clinical research ed.), 388, p.e080952.

Swart, Y. et al., 2021. Local ancestry adjusted Allelic association analysis robustly captures tuberculosis susceptibility loci. Frontiers in genetics, 12, p.716558.

Tremblay, J. et al., 2021. Polygenic risk scores predict diabetes complications and their response to intensive blood pressure and glucose control. Diabetologia, 64(9), pp.2012–2025.

Wang, Y. et al., 2022. Challenges and opportunities for developing more generalizable polygenic risk scores. Annual review of biomedical data science, 5(1), pp.293–320.

